# Enhancing Automated Medical Coding: Evaluating Embedding Models for ICD-10-CM Code Mapping

**DOI:** 10.1101/2024.07.02.24309849

**Authors:** Vanessa Klotzman

## Abstract

**Purpose:** The goal of this study is to enhance automated medical coding (AMC) by evaluating the effectiveness of modern embedding models in capturing semantic similarity and improving the retrieval process for ICD-10-CM code mapping. Achieving consistent and accurate medical coding practices is crucial for effective healthcare management.

**Methods:** We compared the performance of embedding models, including text-embedding-3-large, text-embedding-004, voyage-large-2-instruct, and mistral-embed, against ClinicalBERT. These models were assessed for their ability to capture semantic similarity between long and short ICD-10-CM descriptions and to improve the retrieval process for mapping diagnosis strings from the eICU database to the correct ICD-10-CM codes.

**Results:** The text-embedding-3-large and text-embedding-004 models outperformed ClinicalBERT in capturing semantic similarity, with text-embedding-3-large achieving the highest accuracy. For ICD-10 code retrieval, the voyage-large-2-instruct model demonstrated the best performance. Using the 15 nearest neighbors provided the best results. Increasing the number beyond this did not improve accuracy due to a lack of meaningful information.

**Conclusion:** Modern embedding models significantly outperform specialized models like ClinicalBERT in AMC tasks. These findings underscore the potential of these models to enhance medical coding practices, in spite of the challenges with ambiguous diagnosis descriptions.

## 1 Introduction

The International Classification of Diseases, Tenth Revision, Clinical Modification (ICD-10-CM)[1] is a standardized system for coding diseases and medical conditions. It includes codes for diseases, symptoms, abnormal findings, complaints, social circumstances, and external causes of injury or diseases, providing a common language for recording, reporting, and monitoring diseases. This system is widely used by healthcare providers in the United States as it aids in the classification and coding of diagnoses, symptoms, and procedures. Healthcare organizations often hire medical coders for this task, which can be expensive, time-consuming, and prone to errors. Coding inaccuracies can lead to significant financial implications, contributing to an estimated annual expenditure of 25 billion dollars in the United States[2, 3]. Automatic medical coding (AMC) can alleviate this burden. AMC [4] refers to the automation of clinical coding using machine learning techniques. Despite significant progress in AMC-related work [5–11], challenges persist due to the complexity of extracting knowledge from patients’ clinical records, complex medical terminologies, and variations in descriptions among physicians [12, 13].

Mullenbach et al. [14] used a Convolutional Neural Network (CNN) with an attention mechanism and they leveraged important text snippets to improve the accuracy and interpretability of medical code predictions. However, this method has limitations due to the embedding models they used, such as word2vec, which may not fully capture clinical language and context. But, with the recent rise of Large Language Models (LLMs) they provided new word, sentence, and document embedding models. While LLMs have shown success in various NLP tasks, it is unclear if their performance improvement is due to their larger scale or if the embeddings they produce are significantly different from classical encoding models [15]. The study aims to determine whether modern embedding models from these LLM providers, without utilizing complex architectures like CNNs, can fully capture clinical language correctly.

In this study, we wutilize the eICU dataset and various embedding models from different Large Language Model (LLM) providers: text-embedding-3-large from OpenAI ^1^, text-embedding-004 from Google ^2^, mistral-embed from Mistral AI ^3^, and voyage-large-2-instruct from Voyage-AI ^4^. Using ClinicalBERT [16] as a baseline, the aim is to determine if these modern embedding models can outperform ClinicalBERT in mapping text snipplets, automated medical code mapping, leading to more consistent and accurate medical coding practices. We will answer the research question: Can embedding models from various LLM providers (OpenAI, Google, Mistral AI, and Voyage-AI) outperform ClinicalBERT in accurately mapping clinical descriptions to ICD-10-CM codes?

The contribution of this study is to evaluate the effectiveness of different modern embedding models from various LLM providers (OpenAI, Google, Mistral AI, and Voyage-AI) in accurately map text snippets of diagnoses to ICD-10-CM codes. This evaluation will help determine whether these models can capture clinical language and context without the need for complex architectures like CNNs.

## 2 Materials and Methods

### 2.0.1 eICU Diagnosis Dataset

We utilized the diagnosis strings from patients in the eICU Collaborative Research Database [17], which consists of patient data from various critical care units (CCUs) across the United States, focusing on patients admitted between 2014 and 2015. The diagnosis strings in this database that we use were created by trained eICU clinicians using the APACHE IV diagnosis system [18]. These strings were developed by combining detailed clinical information about the patient’s condition into comprehensive descriptions.For example, a diagnosis string might be “burns/trauma | trauma - skeletal | bone fracture(s) | thoracic spine,” which indicates that a patient has burns and trauma, trauma to the skeletal system, bone fractures, and injuries to the thoracic spine.

### 2.0.2 ICD-10-CM Code Dataset

Each ICD-10-CM code has an associated long description and short description. The long description of the ICD-10-CM code is a detailed explanation of the diagnosis or condition. The short description is a concise summary of the diagnosis or condition. For example, the long description for the ICD-10-CM code “Z87.768” is “Personal history of (corrected) congenital malformations of integument, limbs, and musculoskeletal system,”. The short description is “Prsnl hx of congen malform of integument, limbs and ms sys.” We used the 2015 International Classification of Diseases, 10th Revision, Clinical Modification (ICD-10-CM). Throughout this experiment, we are using the 2015 edition because it aligns the closet with the time period of the eICU dataset. We collected this data from the ICD-10-CM 2015 code descriptions available on the CMS website ^5^.

### 2.1 Model Selection

We selected text-embedding-3-large from OpenAI, text-embedding-004 from Google, mistral-embed from Mistral AI, and voyage-large-2-instruct from Voyage-AI based on their effective performance in various natural language processing tasks according to the Massive Text Embedding Benchmark (MTEB) ^6^. We did not use Meta’s Llama3 because Meta does not have its own embedding model. We chose ClinicalBERT as a baseline for comparison because its embeddings are tailored to clinical texts, accurately capturing medical terminology and making it a reliable standard for evaluating the semantic similarity and retrieval performance of modern embedding models in the medical field. Also ClinicalBERT was trained on a large set of notes from the MIMIC III database containing electronic health records [19]. This makes it an ideal reference point for evaluating it in comparison to newer and more general-purpose LLM embeddings in the context of medical coding.

### 2.2 Assessing Embedding Models for ICD-10-CM Code Retrieval for eICU Diagnoses

We chose to use diagnosis strings that were considered primary diagnoses from the patient’s hospital records in the eICU dataset. Primary diagnoses are well-documented and accurately reflect the main health issues that led to hospitalization. This filtering criteria lead us to then have a dataset of 577,119 diagnosis strings. To enhance the quality of data for our embedding models, we then filtered the dataset to include only those with ICD-9-CM codes successfully mapped to ICD-10-CM codes.This criterion reduced our dataset to only 3,442 diagnoses strings. From these, we random sampled 346 distinct diagnoses without replacement, with a 95% confidence level and a 5% margin of error. Random sampling ensures the sample is unbiased and representative, allowing for accurate and generalizable results. We embedded the models we decided upon with the long descriptions of the ICD-10-CM codes because these descriptions avoid medical abbreviations and capture detailed context, resulting in more accurate and meaningful embeddings. Subsequently, we tested these embedded models using the set of diagnosis strings collected from the eICU database to determine if they could accurately retrieve each diagnosis string’s appropriate ICD-10-CM code. For instance, if we test the embedding model with the diagnosis string “burns/trauma | trauma - CNS | spinal cord injury | cervical,” which is associated with the ICD-10-CM code S14.1, the model should accurately retrieve this code as one of the most similar associations. We tested the embedding models by tweaking the number of nearest neighbors until the results stopped changing significantly.

## 3 Results

The results are presented in Table 1 and highlight the performance of five different models: ClinicalBERT, text-embedding-004, voyage-large-2-instruct, Mistral, and text-embedding-3-large. We assessed the models’ accuracy in matching the correct ICD-10-CM codes to the diagnosis strings created by trained eICU clinicians using the APACHE IV diagnosis system. To determine if there is a significant association between the model type and the correctness of predictions, we conducted a Chi-Square Test for Independence. This test evaluates whether the correctness of predictions is related to the type of model used.

**Table 1.** Contingency Table for Matching ICD-10 Codes to Diagnosis Strings.

| Model | Correct | Incorrect |
| --- | --- | --- |
| voyage-large-2-instruct | 326 | 20 |
| text-embedding-3-large | 299 | 47 |
| text-embedding-004 | 296 | 50 |
| ClinicalBERT | 30 | 316 |
| Mistral | 15 | 331 |

The hypotheses for this test are:

- **Null Hypothesis (H**_0_): There is no association between the type of model and the correctness of predictions.
- **Alternative Hypothesis (H**_1_): There is an association between the type of model and the correctness of predictions.

The categorical variables in this analysis are **Model Type**, representing the five different models being evaluated and **Correctness of Predictions**, indicating whether the predictions made by each model are *correct* or *incorrect*. The observed frequencies of correct and incorrect predictions for each model are presented in the contingency Table 1.

The expected frequencies were calculated for each cell, and the Chi-Square statistic was computed as follows:

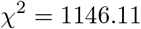

With 4 degrees of freedom, the p-value obtained was extremely small (7.67 × 10^*−*247^), indicating a highly significant result. Since the p-value is much smaller than the typical significance level of 0.05, we reject the null hypothesis (H_0_). We found that the type of model we use affects how often it makes the correct predictions. This means that some models are better at matching ICD-10 codes to diagnosis descriptions than others.

Among the models tested, certain embedding models, specifically voyage-large-2-instruct, text-embedding-3-large, and text-embedding-004, demonstrated superior performance in capturing semantic similarity and enhancing the retrieval process for automated medical code mapping. These improvements resulted in more consistent and accurate medical coding practices. Specifically, using the 15 nearest neighbors for embedding vectors provided the best performance. Increasing the number of neighbors beyond 15 did not improve accuracy. For instance, when matching the diagnosis string “burns/trauma|trauma-head and neck|blunt injury to the neck” to the ground truth ICD-10-CM code S10.8 (Other superficial injuries of neck), including up to 20 neighbors did not enhance accuracy. The additional 5 codes, such as S10.9 (Unspecified superficial injury of neck) and S10.0 (Contusion of neck), were very similar to those in the top 15 and did not provide new information, only adding redundancy.

## 4 Discussion

The results of this study indicate that various embedding models perform differently in automated medical code mapping. Among the models tested, Mistral-embed and ClinicalBERT showed weaker performance, with Mistral-embed struggling even more than ClinicalBERT. Despite ClinicalBERT’s pre-training on biomedical texts and fine-tuning with the Unified Medical Language System (UMLS), it still struggled with specific medical terminologies. The UMLS, a collection of biomedical terms and concepts designed to facilitate the interoperability and integration of diverse health information systems, did not sufficiently enhance ClinicalBERT’s performance. Mistral-embed’s performance was notably poorer, showing significant difficulties with the task.

## 5 Conclusion

This study demonstrates the potential of modern embedding models for improving automated medical code mapping. Models like voyage-large-2-instruct, text-embedding-3-large, and text-embedding-004 outperformed ClinicalBERT and mistral-embed, showing higher accuracy in mapping clinical descriptions to ICD-10-CM codes.

## Data Availability

Data will be available upon request.

## Acknowledgement

I would like to thank the Research Computational Science Team at Children’s Hospital of Orange County for their assistance in clarifying coding systems. Additionally, I appreciate the feedback from the clinical research team and their support in this project.

## Footnotes

1 https://platform.openai.com/docs/guides/embeddings

2 https://ai.google.dev/gemini-api/docs/models/gemini

3 https://mistral.ai/

4 https://www.voyageai.com/

5 https://www.cms.gov/medicare/coding-billing/icd-10-codes

6 https://huggingface.co/spaces/mteb/leaderboard

## References

[1] DiSantostefano, J.: International classification of diseases 10th revision (icd-10). The Journal for Nurse Practitioners 5(1), 56–57 (2009)

[2] Lang, D.: Consultant report-natural language processing in the health care industry. Cincinnati Children’s Hospital Medical Center, Winter 6 (2007)

[3] Farkas, R., Szarvas, G.: Automatic construction of rule-based icd-9-cm coding systems. In: BMC Bioinformatics, vol. 9, pp. 1–9 (2008). Springer

[4] Stanfill, M.H., Williams, M., Fenton, S.H., Jenders, R.A., Hersh, W.R.: A systematic literature review of automated clinical coding and classification systems. Journal of the American Medical Informatics Association 17(6), 646–651 (2010)

[5] Dong, H., Falis, M., Whiteley, W., Alex, B., Matterson, J., Ji, S., Chen, J., Wu, H.: Automated clinical coding: what, why, and where we are? NPJ digital medicine 5(1), 159 (2022)

[6] Teng, F., Ma, Z., Chen, J., Xiao, M., Huang, L.: Automatic medical code assignment via deep learning approach for intelligent healthcare. IEEE journal of biomedical and health informatics 24(9), 2506–2515 (2020)

[7] Mustafa, A., Rahimi Azghadi, M.: Automated machine learning for healthcare and clinical notes analysis. Computers 10(2), 24 (2021)

[8] Zhao, S., Diao, X., Xia, Y., Huo, Y., Cui, M., Wang, Y., Yuan, J., Zhao, W.: Automated icd coding for coronary heart diseases by a deep learning method. Heliyon 9(3) (2023)

[9] López-García, G., Jerez, J.M., Ribelles, N., Alba, E., Veredas, F.J.: Explainable clinical coding with in-domain adapted transformers. Journal of Biomedical Informatics 139, 104323 (2023)

[10] Bhutto, S.R., Zeng, M., Niu, K., Khoso, S., Umar, M., Lalley, G., Li, M.: Automatic icd-10-cm coding via lambda-scaled attention based deep learning model. Methods 222, 19–27 (2024)

[11] Yuan, Z., Tan, C., Huang, S.: Code synonyms do matter: Multiple synonyms matching network for automatic icd coding. arXiv preprint arXiv:2203.01515 (2022)

[12] Yan, C., Fu, X., Liu, X., Zhang, Y., Gao, Y., Wu, J., Li, Q.: A survey of automated international classification of diseases coding: development, challenges, and applications. Intelligent Medicine 2(03), 161–173 (2022)

[13] Ji, S., Sun, W., Dong, H., Wu, H., Marttinen, P.: A unified review of deep learning for automated medical coding. arXiv preprint arXiv:2201.02797 (2022)

[14] Mullenbach, J., Wiegreffe, S., Duke, J., Sun, J., Eisenstein, J.: Explainable prediction of medical codes from clinical text. arXiv preprint arXiv:1802.05695 (2018)

[15] Freestone, M., Santu, S.K.K.: Word embeddings revisited: Do llms offer something new? arXiv preprint arXiv:2402.11094 (2024)

[16] Alsentzer, E., Murphy, J.R., Boag, W., Weng, W.-H., Jin, D., Naumann, T., McDermott, M.: Publicly available clinical bert embeddings. arXiv preprint arXiv:1904.03323 (2019)

[17] Pollard, T.J., Johnson, A.E., Raffa, J.D., Celi, L.A., Mark, R.G., Badawi, O.: The eicu collaborative research database, a freely available multi-center database for critical care research. Scientific data 5(1), 1–13 (2018)

[18] Zimmerman, J.E., Kramer, A.A., McNair, D.S., Malila, F.M.: Acute physiology and chronic health evaluation (apache) iv: hospital mortality assessment for today’s critically ill patients. Critical care medicine 34(5), 1297–1310 (2006)

[19] Johnson, A.E., Pollard, T.J., Shen, L., Lehman, L.-w.H., Feng, M., Ghassemi, M., Moody, B., Szolovits, P., Anthony Celi, L., Mark, R.G.: Mimic-iii, a freely accessible critical care database. Scientific data 3(1), 1–9 (2016)

